# Diagnosing a cardiac cause for palpitations: Five-year service evaluation of a smartphone ambulatory ECG pathway for Emergency Department patients

**DOI:** 10.64898/2026.11.05.26367104

**Authors:** Cormac Morran, Preeti Srinivasan, Rosie Andrew, Ewan AM Pow, Luisa Padovani, Ysabelle Thackray, Ross Murphy, Goran Zangana, Sean Krupej, Mark I’Anson, Lynn Findlay, James Millar, Freya Watts, Maskura Moni, Matthew J Reed

## Abstract

**Background:** Capturing an electrocardiogram (ECG) during transient symptoms remains challenging in emergency department (ED) patients with palpitations. Following the IPED trial, a smartphone-based ambulatory ECG event recorder pathway was integrated into NHS care. We evaluated this service over five years, assessing diagnostic yield, management, one-year outcomes and predictors of cardiac arrhythmia.

**Methods:** Single-centre service evaluation of consecutive patients referred with palpitations to a smartphone-based ambulatory ECG event recorder clinic between 24^th^ July 2019 and 24^th^ July 2024. Patients underwent event-triggered ECG monitoring using the single-lead Kardia Mobile^®^ (AliveCor, Inc.) or their own smartwatch with ECG capability. Clinical characteristics, rhythm recordings, investigation, one-year outcomes and management changes were recorded. Multivarible logistic regression identified independent predictors of one-year cardiac arrhythmia diagnosis.

**Results:** 1750 patients attended the clinic, mean age 44.3 ± 16.1 years, 58.2% female. 1176 patients (67.2%) underwent monitoring with a cardiac arrhythmia directly captured in 67 patients (5.7% of monitored), including atrial fibrillation (n=39), supraventricular tachycardia (13), atrial flutter (9), atrial tachycardia (5) and ventricular tachycardia (1). By one year, 84 (7.1%) who underwent monitoring received a cardiac diagnosis with 79.8% (67/84) established directly via clinic monitoring. Across the entire cohort (n=1750), 114 (6.5%) had a cardiac cause at one year, 1472 (84.2%) had non-cardiac aetiology and 1569 (89.1%) were discharged back to primary care. Medication changes were initiated in 106 patients (6.1%). Independent predictors of a one-year cardiac diagnosis were increasing age (aOR 1.04 per 1-year increase), prior atrial fibrillation/flutter (aOR 5.57) and a presenting symptom of ‘irregular beating’ (aOR 2.65).

**Conclusion:** Our service provides an effective model for evaluating ED patients with palpitations. Over five years the service reliably detected actionable cardiac arrhythmias while safely discharging nearly 90% of patients to primary care. Clinical characteristics may guide patient selection to maximise clinic efficiency.

**Key messages:** *What is already known on this topic:* Although the IPED trial established that smartphone-based ambulatory ECG event recorders improve diagnostic yield for emergency department (ED) patients with palpitations over standard care, data on long-term real-world performance, safety and baseline diagnostic predictors are lacking.

*What this study adds:* In a five-year evaluation of 1750 ED and acute medical unit patients, smartphone-based ambulatory ECG event recorder monitoring directly established nearly 80% of all 1-year cardiac arrhythmia diagnoses while safely discharging 89% of attendees back to primary care without follow-up. Advancing age, prior atrial fibrillation or flutter, and presenting with ‘irregular beating’ were independent predictors of a 1-year cardiac diagnosis.

*How this study might affect research, practice, or policy:* A smartphone-based ambulatory ECG event recorder clinic is an effective model for evaluating patients with palpitations. Emergency, acute and primary care clinicians can use baseline clinical predictors such as increasing age, cardiac history and a description of irregular beating to prioritise those patients more likely to have an underlying cardiac cause for their palpitations for monitoring and effectively guide local service referral strategies.

## INTRODUCTION

Palpitations account for over 300,000 emergency department (ED) visits annually in the UK, representing a major diagnostic challenge for emergency and acute care clinicians. **[1]** While the vast majority are due to benign non-cardiac conditions (i.e. sinus tachycardia or ectopic beats), underlying cardiac arrhythmias including atrial fibrillation (AF) and supraventricular tachycardia (SVT) reduce quality of life, carry risks of stroke and heart failure, and extremely rarely, sudden cardiac death, if unrecognised.

The diagnostic gold standard requires symptom/rhythm correlation by capturing an ECG during symptoms. However, because palpitations are typically intermittent, the presenting ED 12-lead ECG is commonly normal and frequently fails to establish the underlying rhythm diagnosis. Traditional secondary monitoring pathways such as 24- and 48-hour Holter monitors have diagnostic capture rates as low as 10-15% **[2]** due to limited monitoring duration, low symptom frequency and long waits for investigation due to NHS service demands.

The Investigation of Palpitations in the ED (IPED) randomised controlled trial, **[3]** demonstrated that patient-activated, smartphone-based ambulatory ECG event recorder devices markedly increased diagnostic yield (over five-fold) and reduced time to diagnosis by almost 80% compared to standard care. Following the IPED trial, NHS Lothian established a dedicated smartphone palpitations service in the ambulatory care outpatient department at the Royal Infirmary of Edinburgh as part of routine acute care pathways. While clinical trials provide rigorous efficacy data, evaluating long-term real-world implementation is crucial to determine service sustainability, health system impact, safety and diagnostic utility across unselected clinical populations.

This service evaluation evaluates the five-year performance of a real-world smartphone-based ambulatory ECG event recorder palpitations service, examining diagnostic yield, immediate management changes, one-year clinical outcomes and baseline clinical predictors of underlying cardiac arrhythmia.

## METHODS

### Study design and setting

Single-centre service evaluation study at the Royal Infirmary of Edinburgh, a United Kingdom (UK) tertiary academic hospital receiving over 120,000 annual emergency department (ED) visits. The study evaluated a consecutive cohort of patients referred from the ED and Acute Medical Unit (AMU) to the post-ED smartphone palpitations service between 24^th^ July 2019 and 24^th^ July 2024. The study was deemed by the Health Research Authority (HRA; https://www.hra-decisiontools.org.uk/research/) to not require HRA ethical approval. The study was registered as a service evaluation on the NHS Lothian Emergency Department Quality Improvement Project database.

### Patient population

Patients aged 16 years or older presenting to the ED or AMU with acute palpitations in whom initial ED evaluation (including history, physical examination, and baseline 12-lead ECG) revealed no underlying cause or reason for immediate inpatient admission, were eligible for referral. **[4]**

### Clinical pathway

Referred patients attended a next working day outpatient appointment where the ambulatory clinic team (made up of acute medicine clinicians and nurses) evaluated symptom characteristics and supplied the patient with a single-lead smartphone ECG event recorder device (KardiaMobile^®^, AliveCor, Inc., Mountain View, CA, USA) unless their own smartwatch possessed ECG functionality. **[5]** Patients were instructed how to record a 30-second single-lead ECG trace whenever they experienced palpitations over a 7-to-14-day monitoring window. Recordings were reviewed by the ambulatory clinic team at a subsequent clinic follow-up. Specific details about the smartphone palpitations service have previously been reported **[6]** and clinic pathways are available. **[4]**

### Data collection

Clinical data was prospectively recorded the Electronic Patient Record (EPR) and subsequently on a dedicated secure REDCap database hosted at the University of Edinburgh. **[7,8]** Data collected included:

1. *Baseline demographics and clinical features*: Age, sex, primary presenting symptom, symptom duration and past medical history (hypertension, ischemic heart disease, AF/flutter, valvular disease).
2. *Clinic diagnostic yield*: Specific rhythm recorded on the smartphone device during symptomatic episodes.
3. *Immediate clinical management*: Medication changes initiated at clinic (e.g. beta-blockers, anticoagulation, antiarrhythmics) and secondary investigations ordered (e.g. echocardiogram, extended Holter).
4. *One-year outcomes*: Final diagnostic classification at one year (Cardiac cause versus non-cardiac cause versus undiagnosed), specialist cardiology referrals, one-year ED re-attendances, and hospital admissions related to palpitations.

### Primary outcome

The primary outcome was a confirmed cardiac diagnosis at one year, defined as documented atrial fibrillation, atrial flutter, atrial tachycardia, supraventricular tachycardia (SVT; including undifferentiated narrow-complex tachycardia) or ventricular tachycardia. Non-cardiac diagnoses included sinus rhythm, sinus tachycardia, sinus bradycardia, isolated ectopic beats and bigeminy.

### Statistical Analysis

Continuous variables were summarized using means ± standard deviations and categorical variables were expressed as frequencies and percentages. Analyses were performed using available data with no imputation of missing values. Univariable logistic regression screening was performed on all baseline candidate predictors available at initial presentation (n=1176 monitored cohort). Candidate predictors demonstrating p<0.10 in univariable screening, alongside key demographic controls (sex), were entered into a multivariable logistic regression model. Automated backward stepwise elimination based on p<0.05 was conducted to identify independent baseline predictors of a one-year cardiac diagnosis. Model performance was evaluated using the Area Under the Receiver Operating Characteristic curve (AUC). Statistical analyses were performed using Python 3.10 in Google Gemini 3.6 Flash.

## RESULTS

During the five-year study period, 1750 consecutive patients attended the smartphone ambulatory clinic **[Figure 1]**.

**Figure 1.**
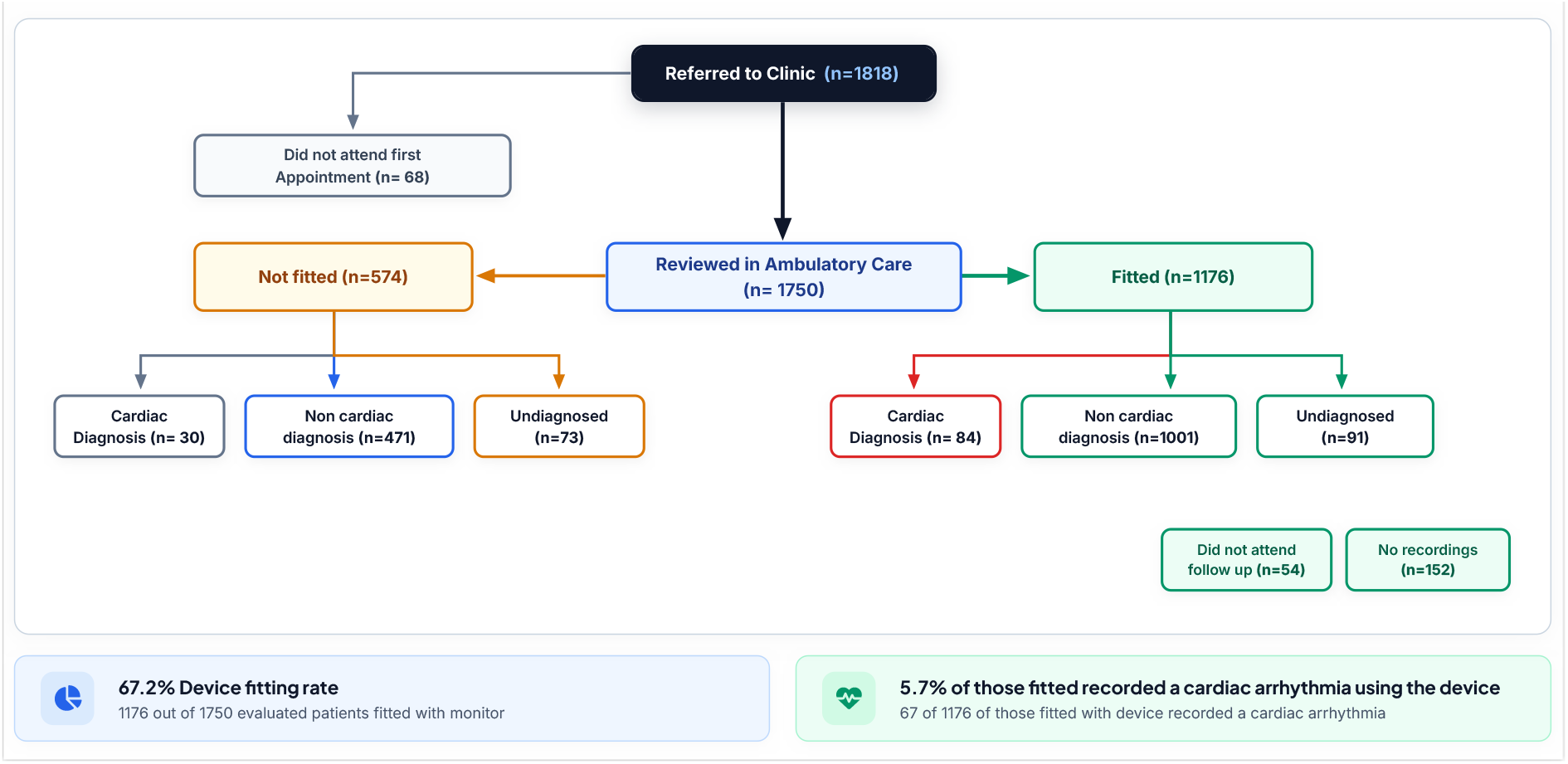
Flow of patients through clinic process

The mean age was 44.3 ± 16.1 years (range 16–93) and 1012 (58.2%) were female. Overall, 1176 (67.2%) patients underwent smartphone ECG monitoring. The remaining 574 patients were not appropriate for device fitting (e.g. presented with isolated chest pain or syncope, incompatible phone). The mean device monitoring duration was 7.0 ± 8.6 days (median 7.0). Baseline demographics and clinical presentations are summarised in **Table 1**.

**Table 1.**
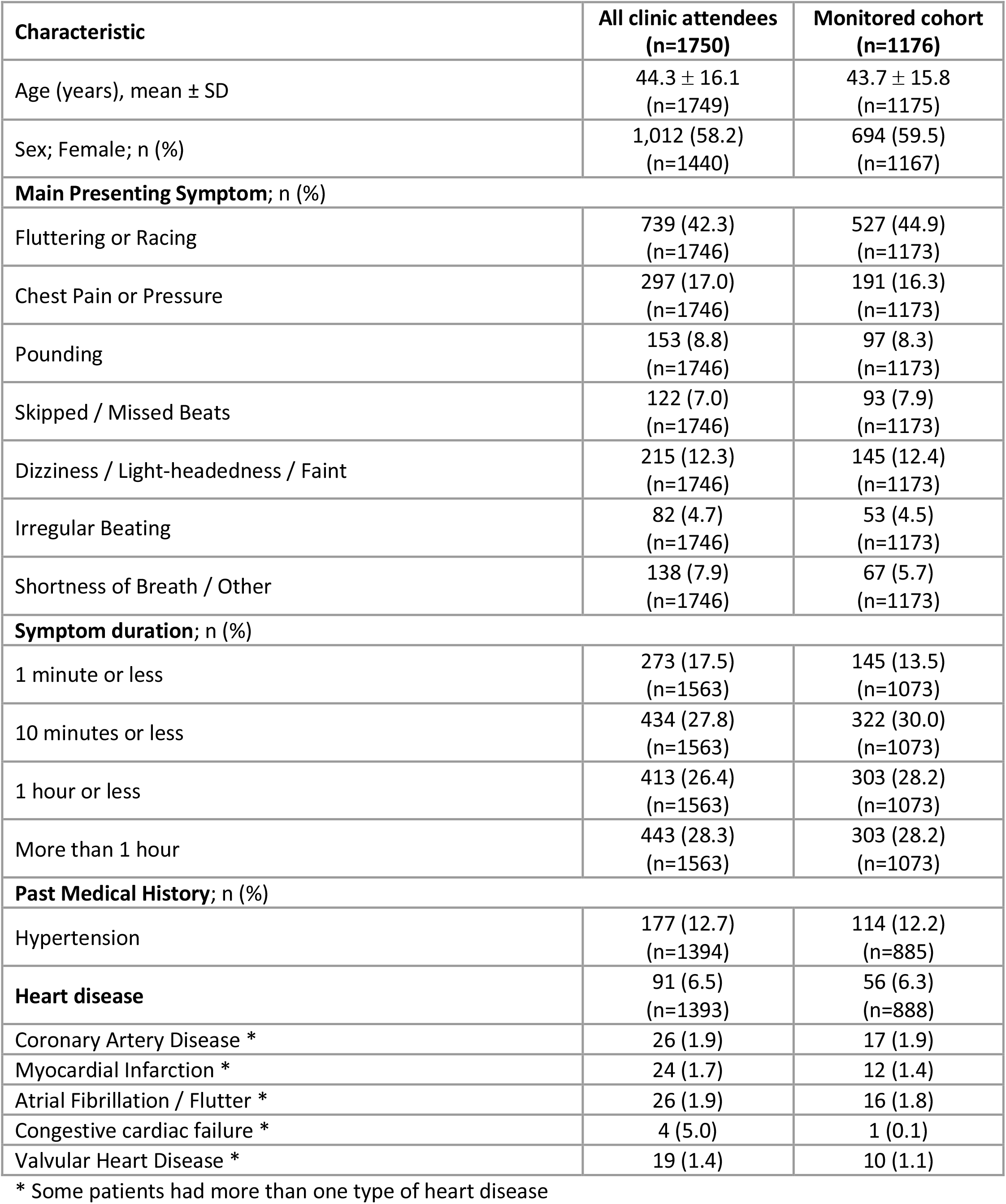
Baseline demographics and presentation characteristics of the study cohort.

| **Characteristic** | **All clinic attendees (n=1750)** | **Monitored cohort (n=1176)** |
|---|---|---|
| Age (years), mean ± SD | 44.3 ± 16.1<br>(n=1749) | 43.7 ± 15.8<br>(n=1175) |
| Sex; Female; n (%) | 1,012 (58.2)<br>(n=1440) | 694 (59.5)<br>(n=1167) |
| **Main Presenting Symptom**; n (%) |  |  |
| Fluttering or Racing | 739 (42.3)<br>(n=1746) | 527 (44.9)<br>(n=1173) |
| Chest Pain or Pressure | 297 (17.0)<br>(n=1746) | 191 (16.3)<br>(n=1173) |
| Pounding | 153 (8.8)<br>(n=1746) | 97 (8.3)<br>(n=1173) |
| Skipped / Missed Beats | 122 (7.0)<br>(n=1746) | 93 (7.9)<br>(n=1173) |
| Dizziness / Light-headedness / Faint | 215 (12.3)<br>(n=1746) | 145 (12.4)<br>(n=1173) |
| Irregular Beating | 82 (4.7)<br>(n=1746) | 53 (4.5)<br>(n=1173) |
| Shortness of Breath / Other | 138 (7.9)<br>(n=1746) | 67 (5.7)<br>(n=1173) |
| **Symptom duration**; n (%) |  |  |
| 1 minute or less | 273 (17.5)<br>(n=1563) | 145 (13.5)<br>(n=1073) |
| 10 minutes or less | 434 (27.8)<br>(n=1563) | 322 (30.0)<br>(n=1073) |
| 1 hour or less | 413 (26.4)<br>(n=1563) | 303 (28.2)<br>(n=1073) |
| More than 1 hour | 443 (28.3)<br>(n=1563) | 303 (28.2)<br>(n=1073) |
| **Past Medical History**; n (%) |  |  |
| Hypertension | 177 (12.7)<br>(n=1394) | 114 (12.2)<br>(n=885) |
| **Heart disease** | 91 (6.5)<br>(n=1393) | 56 (6.3)<br>(n=888) |
| Coronary Artery Disease * | 26 (1.9) | 17 (1.9) |
| Myocardial Infarction * | 24 (1.7) | 12 (1.4) |
| Atrial Fibrillation / Flutter * | 26 (1.9) | 16 (1.8) |
| Congestive cardiac failure * | 4 (5.0) | 1 (0.1) |
| Valvular Heart Disease * | 19 (1.4) | 10 (1.1) |

\* Some patients had more than one type of heart disease

Direct event-triggered smartphone ECG recordings captured a cardiac arrhythmia in 67 patients (5.7% of the monitored cohort). Atrial fibrillation was the most common detected arrhythmia (n=39), followed by SVT (13), atrial flutter (9), atrial tachycardia (5) and ventricular tachycardia (1) **[Table 2]**.

**Table 2.** Symptom-triggered smartphone rhythm findings (n=1176; monitored group)

| Recorded rhythm category | n | Percentage of monitored (%) |
|---|---|---|
| **Cardiac arrhythmia** | **67** | **5.7%** |
| Atrial Fibrillation | 39 | 3.3% |
| Supraventricular Tachycardia (SVT) | 13 | 1.1% |
| Atrial Flutter | 9 | 0.8% |
| Atrial Tachycardia | 5 | 0.4% |
| Ventricular Tachycardia | 1 | 0.1% |
| **Non-cardiac rhythms** | **1267** |  |
| Sinus Rhythm * | 693 | 58.9% |
| Sinus Tachycardia (>100 bpm) * | 274 | 23.3% |
| Ectopics / Bigeminy * | 251 | 21.3% |
| Sinus Bradycardia (<40 bpm) * | 16 | 1.4% |
| Other non-cardiac rhythms * | 33 | 2.8% |
| **Non-diagnostic** | **206** |  |
| No recordings submitted during monitoring | 152 | 12.9% |
| Did not attend follow-up appointment | 54 | 4.6% |

\* Some patients had more than one non-cardiac rhythm detected on the symptom-triggered smartphone

In contrast, 1227 benign non-cardiac rhythms were documented during symptomatic episodes in 889 patients, comprising sinus rhythm (693), sinus tachycardia (274), ectopic beats /bigeminy (251), sinus bradycardia (16) and other non-cardiac rhythms (33). 152 patients (12.9%) recorded no rhythm/symptom correlation ECGs during their monitoring period and 54 (4.6%) did not attend their follow-up appointment.

Immediate therapeutic management changes were instituted directly at the clinic for 106 patients (6.1%) across the overall cohort. Among the 92 patients who had changes to or commencement of medication, initiation of beta-blockers occurred in 57 patients (62.0%) and oral anticoagulation was started in 13 patients (14.1%). Other changes to medication included dose modifications (19; 20.7%) and commencement of proton pump inhibitors (9; 9.8%), ACE inhibitors (3; 3.3%), amiodarone (1; 1.1%) and digoxin (1; 1.1%).

Following clinic assessment, additional diagnostic investigations were requested in 341 patients (19.5%), predominantly arranged by the ambulatory clinic medical team (287; 84.2%). These included 24-hour (223; 12.8%), 48-hour (35; 2.0%) or 7-day Holter monitors (7;0.4%), 144 transthoracic echocardiograms (8.2%) and one implantable loop recorder (0.1%). At one-year follow-up across the entire cohort (n=1750), a cardiac diagnosis was established in 114 patients (6.5%). Within the monitored cohort (n=1176), 84 patients (7.1%) received a final one-year cardiac diagnosis, with 79.8% (67/84) of these diagnoses established directly via the clinic smartphone/watch ECG recording. Secondary investigations identified the remaining 17 cardiac cases **[Table 3]**.

**Table 3.** One-year final diagnoses and health resource utilisation.

| **Outcome Measure** | **Total cohort**<br>(n=1750) | **Monitored cohort**<br>(n=1176) |
|---|---|---|
| **1-year final diagnostic category, n (%)** |  |  |
| Non-cardiac cause (Sinus/ Ectopics/ Benign) | 1472 (84.2) | 1001 (85.2) |
| Cardiac cause | 114 (6.5) | 84 (7.1) |
| Undiagnosed | 164 (9.3) | 91 (7.7) |
| **Mechanism of arrhythmia capture** |  |  |
| Direct smartphone/watch ECG recording capture | 67 (58.7) | 67 (79.7) |
| Subsequent testing | 47 (41.3) | 17 (20.3) |
| **Management & disposition, n (%)** |  |  |
| Discharged to routine primary care (no OP appt) | 1569 (89.1)<br>(n=1738) | 1045 (89.2)<br>(n=1775) |
| Outpatient general cardiology follow-up | 151 (8.7)<br>(n=1738) | 114 (9.7)<br>(n=1775) |
| Specialist electrophysiology consultant referral | 18 (1.0)<br>(n=1738) | 16 (1.4)<br>(n=1775) |
| **1-year emergency resource utilisation** |  |  |
| Unplanned ED / AMU re-attendance | 260 (14.9) | 186 (15.8) |
| Hospital admission for palpitations | 39 (2.3) | 22 (1.9) |

Most attendees 1569 (89.1%) were safely discharged to routine primary care without requiring secondary care outpatient follow-up. Specialist outpatient cardiology follow-up was required for 151 patients (8.7%), and 18 patients (1.0%) were referred directly for cardiac electrophysiology evaluation. Over the following one year, 260 patients (14.9%) re-attended the ED or AMU with symptoms, but only 39 (2.3%) required inpatient admission following this attendance. Pick up rate for arrhythmia dropped slightly after the first year of clinic experience but remained stable thereafter **[Table 4]**.

**Table 4.**
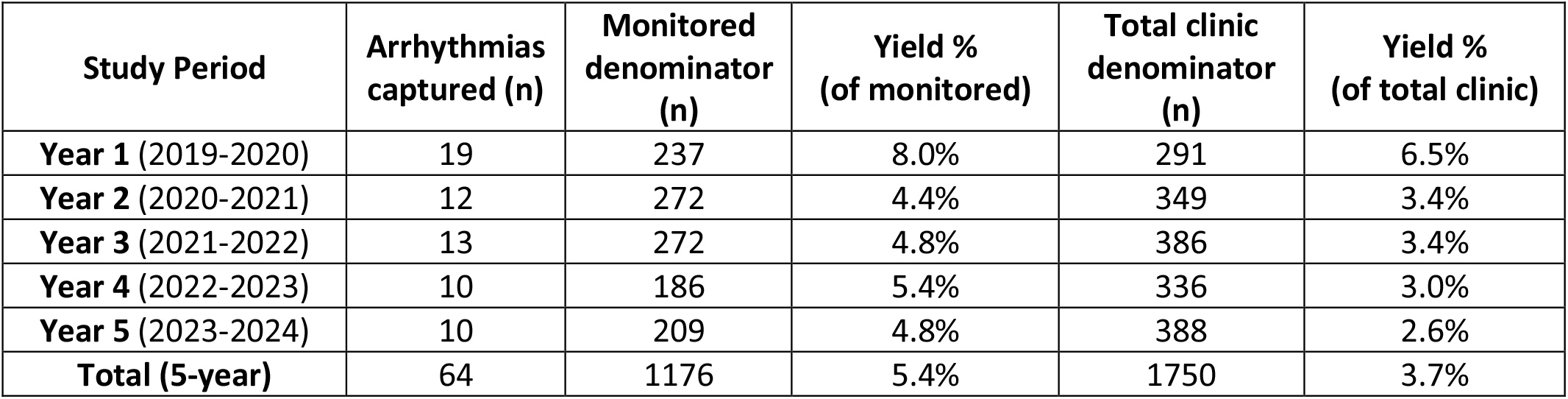
Year-by-year breakdown for smartphone clinic arrhythmia capture.

| **Study Period** | **Arrhythmias captured (n)** | **Monitored denominator (n)** | **Yield %<br>(of monitored)** | **Total clinic denominator (n)** | **Yield %<br>(of total clinic)** |
|---|---|---|---|---|---|
| **Year 1** (2019-2020) | 19 | 237 | 8.0% | 291 | 6.5% |
| **Year 2** (2020-2021) | 12 | 272 | 4.4% | 349 | 3.4% |
| **Year 3** (2021-2022) | 13 | 272 | 4.8% | 386 | 3.4% |
| **Year 4** (2022-2023) | 10 | 186 | 5.4% | 336 | 3.0% |
| **Year 5** (2023-2024) | 10 | 209 | 4.8% | 388 | 2.6% |
| **Total (5-year)** | 64 | 1176 | 5.4% | 1750 | 3.7% |

### Predictors of cardiac arrhythmia

Univariable analysis identified advancing age (OR 1.04, 95%CI 1.03-1.06; p<0.0001), prior history of atrial fibrillation or flutter (OR 8.32, 95%CI 2.95-23.50; p=0.0001), hypertension (OR 3.08, 95%CI 1.73-5.49; p=0.0001), previous cardiac history (OR 2.29, 95%CI 1.05-5.01; p=0.038) and presenting with ‘irregular beating’ (OR 2.86, 95%CI 1.34-6.08; p=0.006) as candidate predictors for a one-year cardiac diagnosis **[Table 5]**.

**Table 5.**
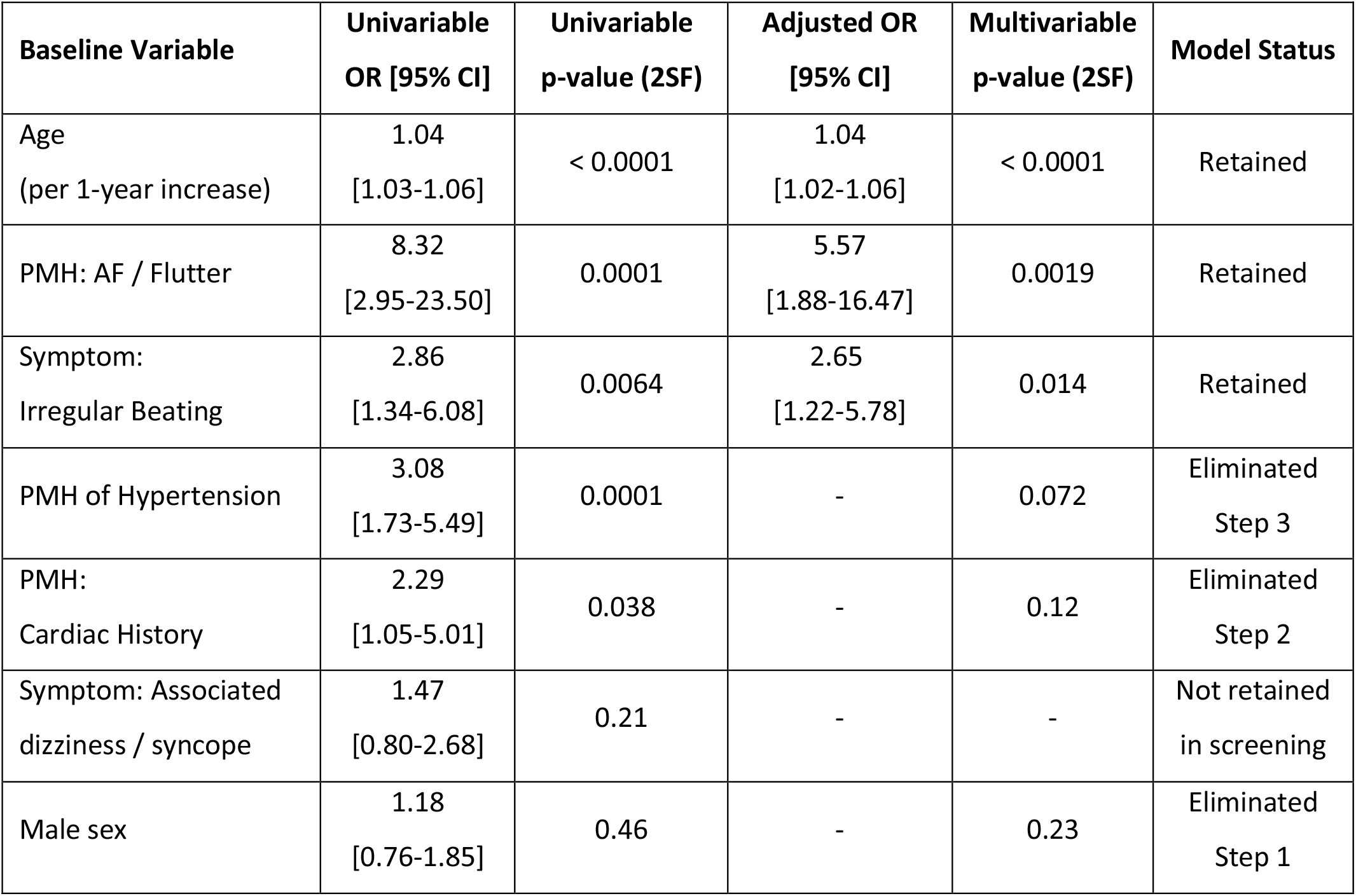
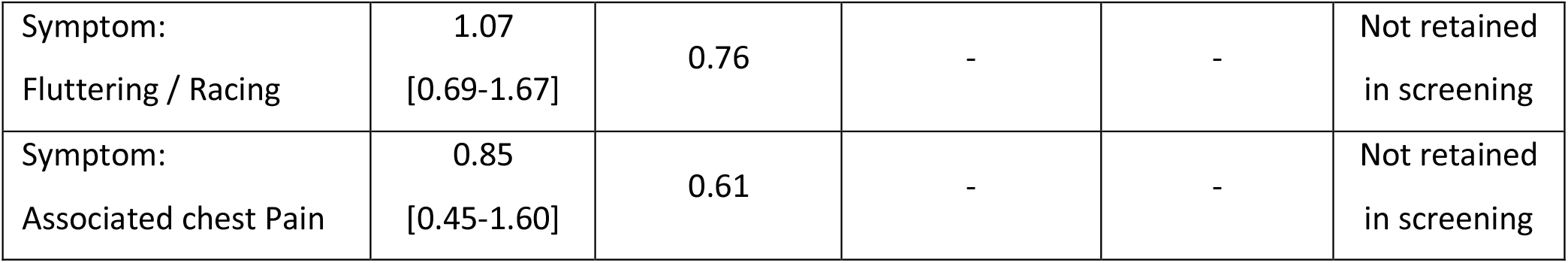
Univariable screening and final backward stepwise multivariable logistic regression model for 1-year cardiac diagnosis (n=84) in the 1176 patients undergoing smartphone monitoring.

Male sex (OR 1.18, 95%CI 0.76-1.85; p=0.46) was included alongside these candidates in the initial multivariable model. Following automated backward stepwise elimination (p<0.05), three baseline clinical features remained significantly associated with a one-year cardiac diagnosis: Age; each one year increase in age increased the odds of a cardiac diagnosis by 4% (adjusted Odd Ratio; aOR 1.04, 95%CI 1.02-1.05; p<0.0001); prior atrial fibrillation or flutter increased the odds of a cardiac diagnosis over five-fold (aOR 5.57, 95%CI 1.88-16.47; p=0.0019); and a main presenting symptom of ‘irregular beating’ increased the odds nearly three-fold (aOR 2.65, 95%CI 1.22-5.78; p=0.014). Male sex (p=0.23), prior cardiac history (p=0.12) and hypertension (p=0.072) were sequentially eliminated during backward selection. The final multivariable model demonstrated discrimination with an AUC of 0.68 (95%CI 0.61-0.75).

## DISCUSSION

This five-year service evaluation study provides real-world evidence supporting the integration of smartphone-based ambulatory event recorder ECG monitoring into emergency diagnostic pathways. 1750 patients were evaluated, demonstrating that this approach can facilitate rapid diagnosis, enable timely intervention and reduce unnecessary referrals to secondary care cardiology services.

In our cohort, around 7% of monitored patients received a confirmed cardiac diagnosis within one year. Crucially, almost 80% of these diagnoses were captured directly on the patient’s initial smartphone ECG event recording during a symptomatic episode enabling immediate initiation of disease-modifying therapies such as oral anticoagulation for AF and atrial flutter to mitigate stroke risk, and beta-blocker therapy to rate control SVT.

Beyond detecting serious pathology, a major strength of the pathway is its ability to rule out cardiac disease and reassure patients who have presented to the ED with palpitations. Over 84% of patients were definitively diagnosed with benign non-cardiac aetiology (such as sinus tachycardia or ectopic beats during symptoms). Demonstrating a normal sinus rhythm trace during symptoms provides powerful reassurance to patients, breaking the anxiety-palpitation cycle. Consequently nearly 90% of clinic attendees were discharged directly back to primary care without requiring secondary cardiology outpatient appointments, relieving pressure on overstretched NHS services.

As digital health technologies proliferate, clinical pathways must avoid over-testing low-risk cohorts. Balen et al studied 215 patients with palpitations and found four independent risk factors were associated with significant cardiac arrhythmia (age ≥ 50 years; OR=7.6, history of heart disease; OR=4.7, syncope as associated symptom; OR=11 and tachycardia ≥100 bpm; OR=3.8). Our multivariable analysis identified three baseline clinical parameters that strongly predict one-year cardiac arrhythmia detection, these being advancing age, history of AF/flutter and symptoms described by the patients as ‘irregular beating’ (rather than ‘racing’ or ‘pounding’). Patients with syncope were not suitable for our service, but looking at both studies together, perhaps increasing age, a cardiac history and a description of irregular beating may perhaps allow clinicians in ED and primary care to prioritise patients for immediate smartphone ECG event monitoring, maximising diagnostic yield and improving service efficiency. We have since emphasised to our ED and AMU teams that referrals should be limited to patients in whom an underlying cardiac cause is considered possible.

This study has limitations. First, this is service evaluation study conducted at a single centre academic tertiary care centre in Scotland and may not translate to other healthcare systems with differing referral structures or digital access. Secondly, missing data occurred in some baseline fields (although overall missing was <1%) due to the retrospective collection of the prospectively written clinic EPR. Equity of access is also a consideration as not all patients have access to a smartphone and/or the ability to use digital technology especially those older patients who may lack familiarity or those with limited capacity or cognition.

## CONCLUSION

A smartphone-based ambulatory ECG clinic represents an effective and safe model for evaluating ED patients presenting with palpitations. Over five years of real-world experience, the pathway demonstrated high diagnostic yield for actionable arrhythmias while allowing almost 90% of patients to be safely discharged to primary care. Applying clinical predictor models based on age, history and symptom description may be able to further refine patient selection for our digital ambulatory care service.

## Data Availability

All data produced in the present study are available upon reasonable request to the authors

